# RELEASE: REdressing Long-tErm Antidepressant uSE in adults in general practice (RELEASE): A two-arm parallel-group cluster RCT effectiveness-implementation hybrid type-1 study

**DOI:** 10.1101/2024.10.08.24314879

**Authors:** a pragmatic multicentre, superiority, parallel group, controlled study of participants allocated to a multi-strategy intervention vs Usual Care, Katharine Wallis, Maria Donald, Mark Horowitz, Rob Ware, Joshua Byrnes, Joanna Moncrieff, Nicholas Zwar, Ian Scott, Mark Morgan, Christopher Freeman, Neeraj Gill, Nancy Sturman, Adam Geraghty, Riitta Partanen, Johanna Lynch

## Abstract

The REdressing Long-tErm Antidepressant uSE in general practice (RELEASE) trial is a pragmatic cluster randomized controlled trial (cRCT) that seeks to determine whether a novel multi-strategy intervention that supports safe cessation of long-term antidepressants is superior to Usual Care.^1^ Pragmatic trials are designed to determine how well interventions work in clinical practice under everyday conditions.^2^

The multi-strategy intervention has two forms, known as RELEASE and RELEASE+. RELEASE for patients includes direct patient education and resources^3^ and an invitation to medication review; RELEASE for general practitioners (GPs) includes education, training in the brief intervention (Ask, Advise, Assist) and printable resources via practice management software. RELEASE+ additionally includes for patients links to freely available information online via an App, and for GPs clinical audit and feedback on practice prescribing of antidepressants. Patient directed education material has the potential to support safer prescribing, including in this case improved recognition and management of antidepressant withdrawal symptoms.^4, 5^ Brief interventions have the potential to change GP prescribing behaviour.^6^ The primary comparison of interest is whether any RELEASE intervention is superior to Usual Care. A secondary comparison will be between the two types of RELEASE interventions (RELEASE and RELEASE+).

The purpose of this document is to minimise bias and ensure transparency and internal validity for the findings of the trial, by defining and making publicly available the analysis approach prior to reviewing or analysing trial data. The statistical analysis plan (SAP) will inform analysis and reporting of the main effectiveness findings of the trial. It provides a detailed description of the primary and secondary trial outcomes and the methods for statistical comparison.

## 1.0 Study synopsis

The study is a pragmatic multicentre, superiority, parallel group, controlled study. The unit of randomisation is the general practice, while the unit of analysis is the individual patient.^1^

### Study identifiers

- Wallis, Katharine A., Donald, Maria, Horowitz, Mark, Moncrieff, Joanna, Ware, Robert S., Byrnes, Joshua, Thrift, Karen, Cleetus, MaryAnne, Panahi, Idin, Zwar, Nicholas, Morgan, Mark, Freeman, Chris, and Scott, Ian (2023). RELEASE (REdressing Long-tErm Antidepressant uSE): protocol for a three-arm pragmatic cluster randomised controlled trial effectiveness-implementation hybrid type-1 in general practice. Trials 24 (1) 615 1-14. https://doi.org/10.1186/s13063-023-07646-w ^1^
- Australian New Zealand Clinical Trials Registry Number: ACTRN12622001379707p
- Ethical approval: The University of Queensland Human Research Ethics Committee (2022/HE001667)

### Protocol variation

The study was initially designed as a two-arm cRCT (effectiveness-implementation hybrid type 1) testing RELEASE vs Usual Care in 24 general practices. Due to additional funding received after the start of the trial and after two practices had been recruited, in mid 2023 the trial transitioned from a two-arm cRCT to a three-arm cRCT to test both RELEASE and RELEASE+ (a more intensive version of RELEASE) versus Usual Care in 28 general practices.

However, due to delays and hurdles in getting signed agreements for development of the RELEASE+ App, we had to pare-back the App from that originally planned to the extent that we now expect the App to add little to the effectiveness of the RELEASE intervention. Therefore, in mid 2024 we changed the analysis plan from RELEASE v Usual Care and RELEASE+ versus Usual Care to RELEASE and RELEASE+ combined versus Usual Care, reflecting that the primary outcomes of interest are the between-group comparisons of RELEASE and RELEASE+ combined versus Usual Care. Thus, while the hypothesis for three-arm trial was that the RELEASE and RELEASE+ interventions were sufficiently different to justify a three-arm trial, the hypothesis was changed to be that the RELEASE and RELEASE+ interventions are essentially the same and will have the same, or similar, effect. This change occurred after participant enrolment began and has no impact on current participants in the trial.

### 1.1 Study objectives

#### 1.1.1 Primary objectives

- To assess the effectiveness of RELEASE combined (i.e. RELEASE and RELEASE+) compared to Usual Care in supporting safe cessation of long-term antidepressants

#### 1.1.2 Secondary objectives

- Assess the effectiveness of RELEASE+ compared to RELEASE in supporting safe cessation of long-term antidepressants
- Assess the cost-effectiveness of RELEASE, RELEASE+ and Usual Care. Cost-effectiveness will be reported in a separate paper.
- Evaluate implementation of the two multi-strategy interventions (e.g. barriers and enablers and implementation outcomes).

#### 1.1.3 Inclusion criteria

##### GP practice inclusion criteria

- Practice uses *Best Practice* or *Medical Director* software (compatible with recruitment software - TorchRecruit®).
- Practice located in south-east Queensland

##### Patient inclusion criteria

- 18 years or older AND
- Currently on antidepressant medication AND
- On any combination of antidepressant medication with a total duration of longer than 12 months including:
  - **SSRIs**
    ◼ Sertraline (Zoloft, Eleva, Sertra, Setrona)
    ◼ Escitalopram (Lexapro, Loxalate, Cilopam, Esipram, Esitalo, Lexam)
    ◼ Fluoxetine (Prozac, Fluotex, Lovan, Prozet, Zactin)
    ◼ Paroxetine (Aropax, Extine, Paroxo, Paxtine, Roxet, Roxtine)
    ◼ Fluvoxamine (Luvox, Faverin, Movox)
    ◼ Citalopram (Celapram, cipramil, Citalo, Talam)
  - **SNRIs**
    ◼ Venlafaxine (Efexor, Elaxine, Enlafax)
    ◼ Desvenlafaxine (Pristiq, Desfax, Desven)
    ◼ Duloxetine (Cymbalta, Andepra, Deotine, Duloxecor, Dytrex, Tixol)
  - **Other antidepressants**
    ◼ Mirtazapine (Avanza, Axit, Mirtanza, Mirtazon)
    ◼ Vortioxetine (Trintellix and Brintellix)
    ◼ Mianserin (Lumin)
    ◼ Moclobemide (Amira, Clobemix, Aurorix)
    ◼ Reboxetine (Edronax)
    ◼ Agomelatine (Valdoxan, Domion)

#### 1.1.4 Exclusion criteria

##### Patient exclusion criteria

- GP considers the patient not suitable for medication review (for example, recent personal crisis)
- Bipolar disorder
- History of psychotic or obsessive-compulsive disorder
- Currently under care of a psychiatrist
- Current substance use disorder
- Non-psychiatric indication for antidepressant (for example, neuropathic pain)
- Dementia or unable to give informed consent
- Aged care residents

### 1.2 Outcomes

#### 1.2.1 Primary outcome

- *Antidepressant discontinuation at 12-months* post patient allocation, self-reported by patients. Discontinuation is defined as 0mg antidepressant maintained for at least 2 weeks.

#### 1.2.2 Secondary outcomes

- *Proportion achieving 75% reduction at 12-months*. A 75% reduction is likely more achievable for people who have been on antidepressants for many years and has health benefits.30
- *Health-related quality of life:* 12-item short-form health survey (SF-12).
- *Antidepressant side effects*: 12-item Antidepressant Side Effect Checklist (ASEC-12).
- *Wellbeing*: The Warwick Edinburgh Mental Well-being Scale (WEMWBS-14)
- *Withdrawal symptoms:* 15-item Distinctive Antidepressant Withdrawal Scale
- *Emotional numbing:* Emotional Reactivity Numbing Scale: General subscale (ERNS-8).
- *Beliefs about antidepressants:* Beliefs about Medications Questionnaire-Specific.
- *Depressive symptoms:* Patient Health Questionnaire (PHQ9).
- Anxiety symptoms: Generalized Anxiety Disorder measure (GAD7).
- *Aggregated practice level antidepressant prescribing data at baseline and 12-months follow-up*.

#### 1.2.3 Other measures

- *Demographic characteristics*: Age, Gender, Relationship status, Educational level, Employment status, Living arrangements, ATSI, Ethnicity, Household income, Residential location (Postcode)
- *Antidepressant prescriptions including dose*: self-reported, and via GP health record
- *Health and lifestyle:* BMI (weight and height); falls in the past 6 months; co-morbidities (e.g., diabetes); current smoking; alcohol use
- *Health service utilisation:* self-report and via GP health record including GP visits, outpatient department visits, Emergency department presentations, hospitalisations, specialist appointments.

### 1.3 Intervention/s

- **Usual Care:** practices may provide medication review as usual. This is permitted as in both arms of the trial *all prescribing decisions are made as usual by GP and patient together*.
- **RELEASE:** The RELEASE interventions are user-informed multi-modal interventions based in general practice targeting both GPs and patients. The nub of the RELEASE intervention/s is the drug-specific hyperbolic tapering plans which have the following implementation strategies:

| GPs: implementers | Patients: empowered recipients |
| --- | --- |
| 1. Practice outreach training visit<br>- RELEASE 3As brief intervention (Ask, Advise, Assist) (CPD)<br>2. eLearning module (~1hr) (CPD)<br>3. Changes to GP practice management software | 1. Invitation to schedule a medication review with GP (mailout and email)<br>2. Awareness raising (mailout and email)<br>- Brochure: "Stopping Antidepressants"<br>- Family and friend's brochure<br>- Links to information (articles, documentaries videos, eMH resources) |
| - Uploading printable RELEASE resources including drug-specific tapering plans, brochures, and Decision Aid<br>- Adding custom prescriptions for compounded antidepressant mini-dose capsules<br>4. Brief instructional videos (~2 min)<br>5. Clinical audit & Feedback for practice antidepressant prescribing (RELEASE+ only)<br>6. Email prompts to initiate RELEASE 3As brief intervention (every 28 days) | 3. Decision Aid (mailout and email)<br>4. Tapering plan (their antidepressant) (mailout and email)<br>5. SMS prompts to schedule a medication review (every 28 days) |

#### RELEASE has the following guiding principles

- All prescribing decisions are made as usual by the GP and patient together.
- Empowering patients.
- Acceptable and translatable in routine general practice.
- Resources developed to raise awareness, prompt medication review, support decision-making, and guide safe cessation of long-term antidepressants.

The RELEASE+ intervention includes for GPs, an additional outreach visit with clinical audit and feedback on practice antidepressant prescribing data; and for patients, use of an App that includes links to freely available evidence-based information and e-mental health support sites

### 1.4 Setting

Twenty-eight GP practices will be recruited through The University of Queensland’s Practice-based Research Network (UQGP Research) and other networks using email, telephone calls and outreach visits. These practices will be in south-east Queensland.

### 1.5 Randomisation

Randomisation is clustered at the practice level to avoid contamination between intervention and Usual Care arms. Practice randomisation occurred after baseline data collection to avoid bias in GP de-selection of potentially eligible patients. Randomisation was designed assuming a three-arm trial in a 3:2:2 Usual Care:RELEASE:RELEASE+ ratio via a central web-based randomisation service (Griffith University). Practices were stratified by size (>5 / <5 FTE GPs). Randomisation occurred in blocks of size 7 or 14, with block size randomly assigned in a 1:1 ratio. The allocation ratios were chosen with regard to optimising sample size for the primary outcome, as well as the secondary comparison within the RELEASE groups.

History of Study: Due to the additional funding received after the start of the trial and subsequent transition from a single RELEASE intervention to the standard RELEASE and RELEASE+ interventions, the first two practices recruited were randomised and allocated into one of two arms (Usual Care or (standard) RELEASE) and thereafter practices were randomised and allocated into Usual Care, RELEASE or RELEASE+.

### 1.6 Data collection

Participant e-survey data at baseline, 6- and 12-months is collected via the secure research data collection tool REDCap (Research Electronic Data Capture, Vanderbilt, USA). For the baseline data, REDCap generates an automated email to participants with a ‘closed’ link to the baseline e-survey.

For the 6- and 12-months data, practices send an SMS to participants three days before the e-survey due date to foreshadow an email coming from REDCap inviting participants to complete the 6- and 12-month e-surveys. REDCap is programmed to email all participants on the survey due date to invite survey completion. For participants who do not respond to this email invitation, REDCap sends a reminder email on days 7, 14, and 21 following the due date; and an SMS reminder on day 4 and if no response again on day 9. For participants who have not responded, the research team phone to invite participation at two weeks after the due date. REDCap is programmed to only send email and SMS reminders to participants who have not yet completed their survey.

### 1.7 Sample size

To calculate sample size, we assumed 12% of participants in the Usual Care group and 30% of participants in each of the intervention groups will achieve the primary outcome (stopped antidepressants by 12 months). Because the study was designed to involve two comparisons, Usual Care vs RELEASE and Usual Care vs RELEASE+, we designed the study to allocate more participants to the Usual Care arm as this increases the overall efficiency of the trial (that is, for fixed type I and type II errors we require fewer total participants). The ratio chosen for allocation was 3:2:2. Using a two-sided alpha significance level of 0.035 (chosen to facilitate comparison of the two intervention arms against a single control arm) and 90% power then, without accounting for clustering, we would require 142 participants in the Usual Care arm and 95 in each of the intervention arms (332 total).

As this is a cluster RCT, we need to account for the non-independence of observations from individual participants recruited from the same practice. We assume there will be a mean of 20 participants recruited from each practice who will provide data on the primary outcome, and that the within-practice intra-cluster correlation = 0.03, giving a design effect of 1.57. Consequently, we require 522 participants to provide data. To achieve this number, and to account for the possible drop-out of practices once enrolled, we aimed to recruit 28 practices. Assuming 20% attrition, we required 653 participants to enroll and provide baseline data. This requires the 28 practices to recruit an average of 23.3 patients.

If the study is considered as a two-arm trial, and continuing to assume 12% of participants in the Usual Care group achieve the primary outcome, these recruitment figures give 90% power to detect a statistically significant difference (two-tailed alpha=0.05) between the RELEASE combined and Usual Care groups of when at least 26% of the RELEASE combined groups have ceased anti-depressants at 12-months.

## 2.0 Statistical analysis

Author: Maharshi Patel, reviewed by Professor Robert Ware

### 2.1 General principles

Outcomes will be analysed on an intention-to-treat basis. The unit of analysis is the individual patient.

All models that investigate patient-level effects will include the stratification factor practice size as a fixed effect, and GP practice as a random effect. Longitudinal associations will be investigated using analyses that account for multiple observations per participant. Individual analyses will be determined by data structure, for example, three-level hierarchical mixed-effects models with ‘patient’ nested within ‘GP’ nested within ‘GP practice’ where appropriate. With regard to missing data, the structure and pattern of missing data will be assessed and if necessary, a sensitivity analysis undertaken based on imputed data using a multiple imputation model (see below). Data will be stored de-identified.

#### 2.1.1 Presentation of results

Continuous data will be summarized descriptively using either mean and standard deviation (SD) or median and interquartile range (IQR), depending on the distribution of the variable of interest. Categorical data will be presented as frequency and percentage.

#### 2.1.2 Level of significance

A significance level of alpha = 0.05 (two-tail) will be used to evaluate statistical significance for the primary outcome.

#### 2.1.3 Data imputation

All analyses will be presented with observed data. Additionally, missing data will be imputed for any outcome with >5% missingness. Data will be imputed using multiple imputation (see Section 3.5, ‘Sensitivity Analysis’, below).

#### 2.1.4 Statistical software

All estimates will be derived using Stata v13 (StataCorp, College Station, Texas, USA).

### 2.2 Evaluation of demographic and baseline characteristics

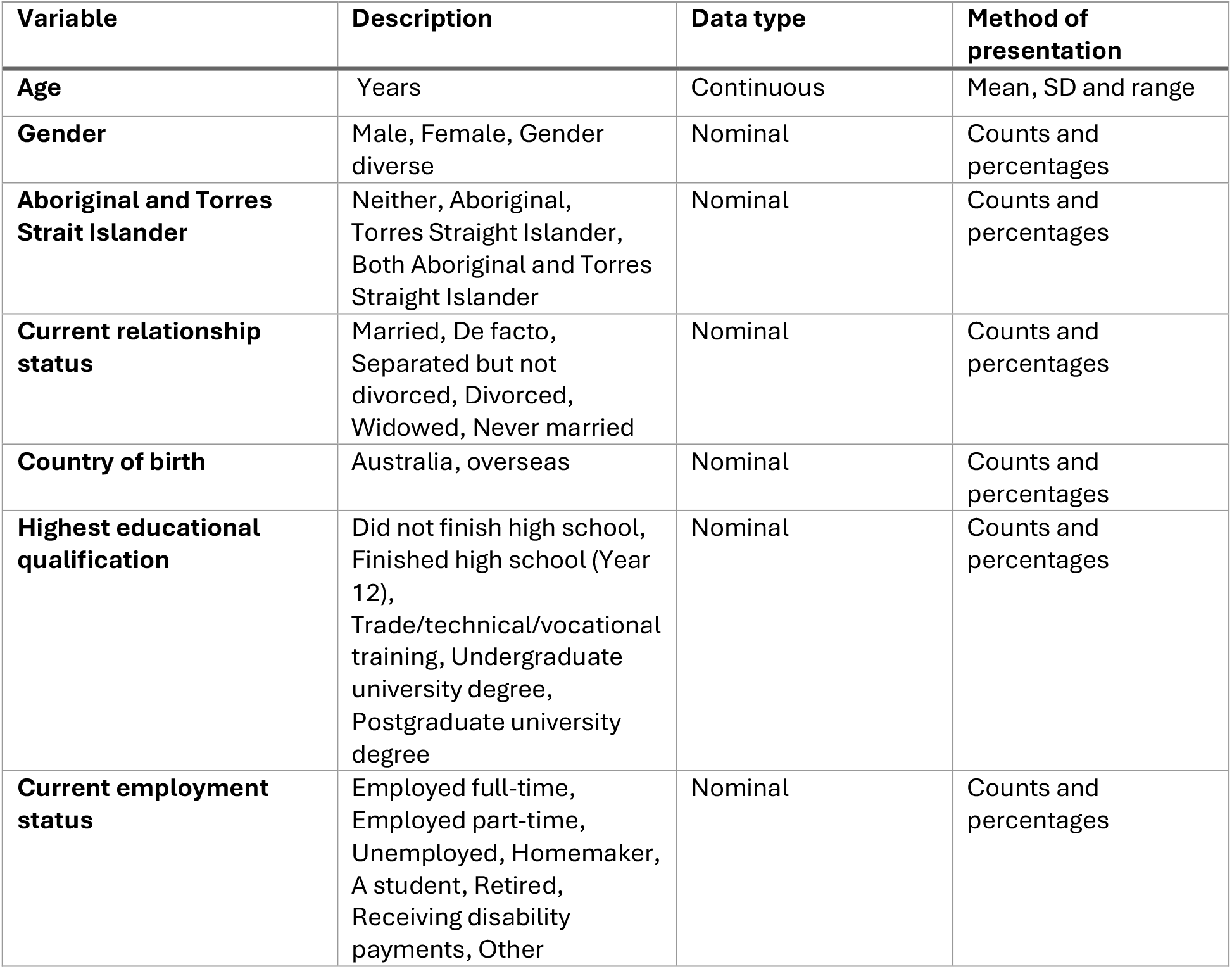
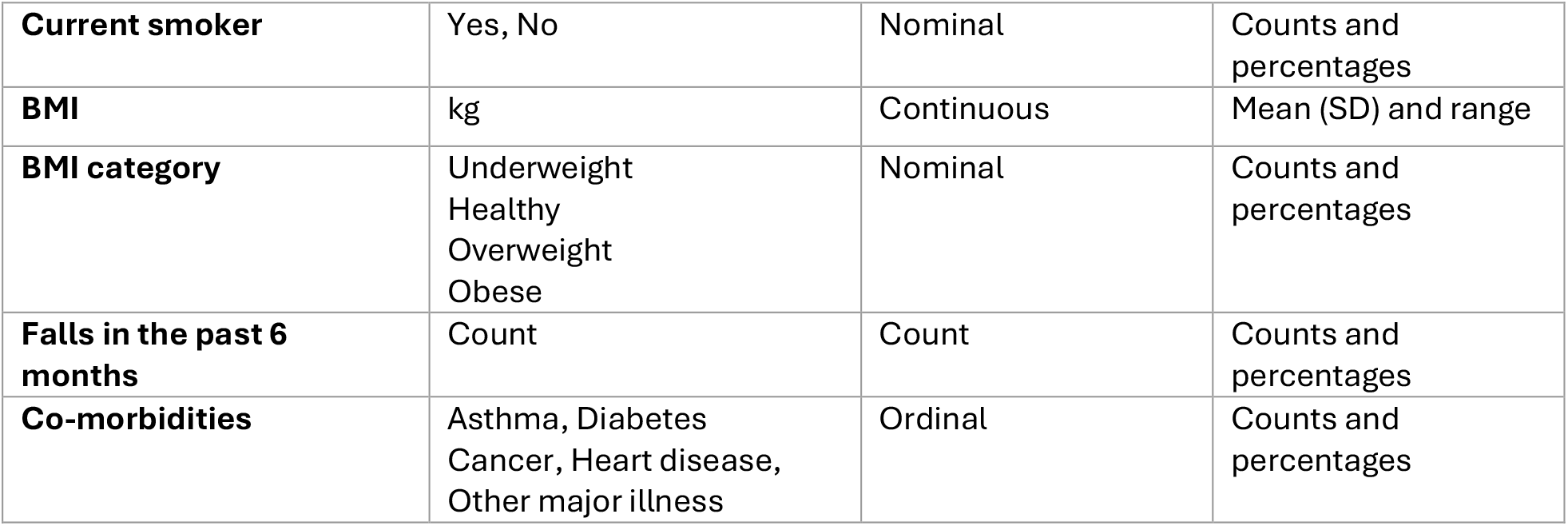

| Variable | Description | Data type | Method of presentation |
| --- | --- | --- | --- |
| **Age** | Years | Continuous | Mean, SD and range |
| **Gender** | Male, Female, Gender diverse | Nominal | Counts and percentages |
| **Aboriginal and Torres Strait Islander** | Neither, Aboriginal, Torres Strait Islander, Both Aboriginal and Torres Strait Islander | Nominal | Counts and percentages |
| **Current relationship status** | Married, De facto, Separated but not divorced, Divorced, Widowed, Never married | Nominal | Counts and percentages |
| **Country of birth** | Australia, overseas | Nominal | Counts and percentages |
| **Highest educational qualification** | Did not finish high school, Finished high school (Year 12), Trade/technical/vocational training, Undergraduate university degree, Postgraduate university degree | Nominal | Counts and percentages |
| **Current employment status** | Employed full-time, Employed part-time, Unemployed, Homemaker, A student, Retired, Receiving disability payments, Other | Nominal | Counts and percentages |
| **Current smoker** | Yes, No | Nominal | Counts and percentages |
| **BMI** | kg | Continuous | Mean (SD) and range |
| **BMI category** | Underweight<br>Healthy<br>Overweight<br>Obese | Nominal | Counts and percentages |
| **Falls in the past 6 months** | Count | Count | Counts and percentages |
| **Co-morbidities** | Asthma, Diabetes<br>Cancer, Heart disease,<br>Other major illness | Ordinal | Counts and percentages |

### 2.3 Planned analysis of the primary outcome

The number and proportion of participants who experience the outcome antidepressant cessation (yes/no) at 12-months. Between-group differences will be assessed using mixed-effects logistic regression, with effect estimates presented as odds ratio (95% CI); *P* value. Treatment group (RELEASE/Usual Care) will be included as the main fixed effect, time will be included as a fixed effect (6-/12-months) along with a group-by-time interaction term, practice size will be included as a fixed effect, and GP practice will be included as a random effect to account for probable non-independence of results from participants who attend the same GP practice.

For the main analysis: participants who actively drop out before assessment will be classified as ‘missing’; participants who are lost to follow-up will be classified as ‘missing’; participants who die during the study will be excluded from analysis.

A supplementary analysis will use a generalised linear model with binomial family and identify link to calculate the absolute risk difference in the primary outcome between the RELEASE and Usual Care groups. This will allow the calculation of the ‘number needed to treat to benefit’ and 95% CI.

### 2.4 Planned analysis of secondary outcomes

Secondary outcomes will also be analysed using mixed-effects models, using linear regression for interval outcomes, logistic regression for binary outcomes, and Poisson regression for count outcomes. When data is recorded at 6- and 12-months mixed-effects models will be used with time included as a main effect and a group-by-time interaction.

For secondary outcomes: participants who actively drop out before assessment will be classified as ‘missing’; participants who are lost to follow-up will be classified as ‘missing’; participants who die during the study will be excluded from analysis.

#### 2.4.1 75% antidepressant dose reduction

The number and proportion of participants who experience the outcome 75% antidepressant dose reduction (yes/no). A 75% reduction is likely more achievable for people who have been on antidepressants for many years and has health benefits.^7^

Between-group differences will be assessed using mixed-effects logistic regression, with effect estimates presented as odds ratio (95% CI); *P* value. Treatment group (RELEASE combined/Usual Care) will be included as the main fixed effect. Time will be included as a fixed effect (6-/12-months) along with a group-by-time interaction term. Practice size will be included as a fixed effect, and GP practice will be included as a random effect.

#### 2.4.2 Health-related quality of life (SF-12)

##### Health-related quality of life

12-item short-form health survey (SF-12).^8^

Rating scales will be treated as continuous variables and the mean and SD calculated per group. A linear regression, with study group as the main effect, will be used to determine the between-group difference. The results will be presented as the mean difference (95% CI). Time will be included as a fixed effect (6-/12-months) along with a group-by-time interaction term. Practice size will be included as a fixed effect, and GP practice will be included as a random effect.

#### 2.4.3 Antidepressant side effects (ASEC-12)

##### Antidepressant side effects

12-item Antidepressant Side Effect Checklist (ASEC-12).^9^

Rating scales will be treated as continuous variables and the mean and SD calculated per group. A linear regression, with study group as the main effect, will be used to determine the between-group difference. Time will be included as a fixed effect (6-/12-months) along with a group-by-time interaction term. Practice size will be included as a fixed effect, and GP practice will be included as a random effect.

#### 2.4.4 Well-being (WEMWBS-14)

##### Wellbeing

The Warwick Edinburgh Mental Well-being Scale (WEMWBS-14).^10^

Rating scales will be treated as continuous variables and the mean and SD calculated per group. A linear regression, with study group as the main effect, will be used to determine the between-group difference. The results will be presented as the mean difference (95% CI). Time will be included as a fixed effect (6-/12-months) along with a group-by-time interaction term. Practice size will be included as a fixed effect, and GP practice will be included as a random effect.

#### 2.4.5 Discriminatory antidepressant withdrawal symptoms (DAWS)

##### Withdrawal symptoms

15-item Distinctive Antidepressant Withdrawal Scale.^11^

Rating scales will be treated as continuous variables and the mean and SD calculated per group. A linear regression, with study group as the main effect, will be used to determine the between-group difference. The results will be presented as the mean difference (95% CI). Time will be included as a fixed effect (6-/12-months) along with a group-by-time interaction term. Practice size will be included as a fixed effect, and GP practice will be included as a random effect.

#### 2.4.6 Emotional numbing (ERNS-General subscale)

##### *Emotional numbing:* Emotional Reactivity Numbing Scale

General subscale (ERNS-8).^12^

Rating scales will be treated as continuous variables and the mean and SD calculated per group. A linear regression, with study group as the main effect, will be used to determine the between-group difference. The results will be presented as the mean difference (95% CI). Time will be included as a fixed effect (6-/12-months) along with a group-by-time interaction term. Practice size will be included as a fixed effect, and GP practice will be included as a random effect.

#### 2.4.7 Beliefs about antidepressants (BMQ-specific)

##### Beliefs about antidepressants

Beliefs about Medications Questionnaire-Specific.^13^

Rating scales will be treated as continuous variables and the mean and SD calculated per group. A linear regression, with study group as the main effect, will be used to determine the between-group difference. The results will be presented as the mean difference (95% CI). Time will be included as a fixed effect (6-/12-months) along with a group-by-time interaction term. Practice size will be included as a fixed effect, and GP practice will be included as a random effect.

#### 2.4.8 Depressive symptoms (PHQ-9)

##### Depressive symptoms

Patient Health Questionnaire (PHQ9).^14^

Rating scales will be treated as continuous variables and the mean and SD calculated per group. A linear regression, with study group as the main effect, will be used to determine the between-group difference. The results will be presented as the mean difference (95% CI). Time will be included as a fixed effect (6-/12-months) along with a group-by-time interaction term. Practice size will be included as a fixed effect, and GP practice will be included as a random effect.

#### 2.4.9 Anxiety symptoms (GAD-7)

##### Anxiety symptoms

Generalized Anxiety Disorder measure (GAD7).^15^

Rating scales will be treated as continuous variables and the mean and SD calculated per group. A linear regression, with study group as the main enect, will be used to determine the between-group dinerence. The results will be presented as the mean dinerence (95% CI). Time will be included as a fixed enect (6-/12-months) along with a group-by-time interaction term. Practice size will be included as a fixed enect, and GP practice will be included as a random enect.

#### 2.4.10 Health and lifestyle: Body Mass Index

Body mass index (BMI) will be treated as a continuous variable and the mean and SD calculated per group. A linear regression, with study group as the main effect, will be used to determine the between-group difference. The results will be presented as the mean difference (95% CI). Time will be included as a fixed effect (6-/12-months) along with a group-by-time interaction term. Practice size will be included as a fixed effect, and GP practice will be included as a random effect.

#### 2.4.11 Health and lifestyle: Falls in the last 6 months

Falls will be treated as a continuous variable and the mean and SD calculated per group. A linear regression, with study group as the main effect, will be used to determine the between-group difference. The results will be presented as the mean difference (95% CI). Time will be included as a fixed effect (6-/12-months) along with a group-by-time interaction term. Practice size will be included as a fixed effect, and GP practice will be included as a random effect.

A supplementary analysis will dichotomise number of falls as ‘any’ or ‘none’ and repeat the analysis using a logistic regression model.

#### 2.4.12 Health and lifestyle: Current smoking

Current smoking will be treated as a binary variable and the frequency and percentage calculated per group. A logistic regression, with study group as the main effect, will be used to determine the between-group difference. The results will be presented as odds ratio (95% CI). Time will be included as a fixed effect (6-/12-months) along with a group-by-time interaction term. Practice size will be included as a fixed effect, and GP practice will be included as a random effect.

#### 2.4.13 Practice level prescribing data

At the practice level, the number and proportion of practice patients on antidepressants, and the number and proportion on long-term antidepressants (longer than 12-months).

Between-group differences will be assessed using mixed-effects linear regression, with effect estimates presented as mean difference (95% CI); *P* value. Treatment group (RELEASE combined/Usual Care) will be included as the main fixed effect Practice size will be included as a fixed effect, and GP practice will be included as a random effect.

If the normality assumption of linear regression is not met, the median and IQR will be calculated per group, and a quantile regression will be performed with study group as the main effect. The results will be presented as the median difference (95% CI).

#### 2.4.14 Health utilisation data

For participants, for the 12-months before and after allocation, data will be collected from the practice electronic health record for GP visits, including number, type (face to face or Telehealth) and length (minutes); and for Mental Health Care plan (yes/no) and Mental Health Plan review (yes/no) for the health economics evaluation.

### 2.5 Sensitivity Analyses

A sensitivity analysis will be conducted to assess any effect of the first two practices randomised prior to the transition to the three-arm trial.

Sensitivity analyses will be conducted for all outcomes to assess the effect of loss to follow up and withdrawal. Multiple imputation by chained equations will be used for all primary and secondary outcomes. Imputation variables will be determined by considering the baseline demographic and clinical characteristics of participants who do, and do not, provide primary outcome data. Variables that are imbalanced will be included as imputation variables. Each outcome will be imputed 50-times.

## 3.0 Trial status

Practice recruitment complete (26 practices). Participant recruitment and baseline data collection will be closed on Sept 21^st^.

Rolling intervention delivery and follow-up data collection is ongoing. Data collection is expected to be complete by November 2025.

## Data Availability

All data produced in the present study are available upon reasonable request to the authors

